# Alzheimer’s disease neuroimaging signature aids identification of cognitive impairment in older adults with early-onset epilepsy

**DOI:** 10.64898/2026.06.05.26354952

**Authors:** McKenna E. Williams, Kayela Arrotta, Katherine J. Bangen, Anny Reyes, Alena Stasenko, Ifrah Zawar, Vineet Punia, Irene Wang, Wanyong Shin, Ting-Yu Su, Jerry J. Shih, Nikdokht Farid, Jaideep Kapur, Aaron F. Struck, Vicente J. Iragui-Madoz, Lynn M. Bekris, Lisa Ferguson, Dace N. Almane, Jana E. Jones, Bruce P. Hermann, Robyn M. Busch, Carrie R. McDonald, the Alzheimer’s Disease Neuroimaging Initiative

**Affiliations:** Department of Neurosciences, University of California San Diego, La Jolla, CA, USA; Epilepsy Center and Department of Neurology, Neurological Institute, Cleveland Clinic, Cleveland, OH, USA; Department of Psychiatry, University of California San Diego, La Jolla, CA, USA; Psychology Service, VA San Diego Healthcare System, San Diego, CA, USA; Department of Neurology, University of Virginia School of Medicine, Charlottesville, VA, USA; Department of Biomedical Engineering, Cleveland Clinic Research, Cleveland, OH, USA; Department of Diagnostic Radiology, Cleveland Clinic, Cleveland, OH, USA; Department of Radiology, University of California San Diego, La Jolla, CA, USA; Department of Neurology, Washington University in St. Louis, St. Louis, MO, USA; Department of Laboratory Medicine and Pathology, University of Washington, Seattle, WA, USA; Department of Neurology, University of Wisconsin School of Medicine and Public Health, Madison, WI, USA

## Abstract

**Background and Objectives:** Older adults with epilepsy are at increased risk for Alzheimer’s disease (AD), yet the mechanisms underlying this association remain poorly understood. We applied a validated AD neuroimaging signature to older adults with epilepsy to examine 1) whether older adults with epilepsy mirror AD-related changes, 2) associations with clinical, cognitive, and plasma biomarker outcomes, and 3) utility for identifying subgroups at heightened risk for cognitive decline. Our multicenter, prospectively enrolled cohort allowed for direct examination of differences in AD signatures between those with early-onset and late-onset unexplained epilepsy.

**Methods:** Participants included 449 older adults: 87 with focal epilepsy from the multicenter Brain Aging and Cognition in Epilepsy (BrACE) cohort (age=66.10 [SD=6.86], including early-onset (<55 years at seizure onset) and late-onset (≥55 years at seizure onset) epilepsy); 362 from the Alzheimer’s Disease Neuroimaging Initiative (ADNI), including cognitively unimpaired (CU) healthy controls and individuals with mild cognitive impairment (MCI) or AD dementia. An AD signature was derived from regional cortical thickness and hippocampal volume weighted by their sensitivity to AD-related neurodegeneration in prior work. Associations between the AD signature, epilepsy characteristics, plasma biomarkers (β-amyloid 42/40, phosphorylated tau [pTau217, pTau181], neurofilament light chain [NfL]), and cognition were evaluated in BrACE.

**Results:** Participants with epilepsy demonstrated more AD-like signatures compared to ADNI CU controls (β= −0.43, *p*_adj_<0.001), reflecting reduced thickness/volume in AD-vulnerable regions. This effect was stronger among early-onset (β= −0.57) versus late-onset (β= −0.26) epilepsy. In BrACE, the AD signature correlated with NfL (β= −0.30, *p*_adj_=0.050), memory performance (β= 0.30, *p*_adj_=0.006), and predicted greater odds of cognitive impairment specifically among those with early-onset, but not late-onset, epilepsy (interaction *p*_adj_=0.043). Further, among those with early-onset epilepsy, the AD signature significantly improved identification of cognitive impairment over and beyond the effects of plasma AD biomarkers (*p*=0.041). Findings were similar when examining the effects of epilepsy duration rather than epilepsy onset age.

**Discussion:** AD neuroimaging signatures may help identify clinically meaningful subgroups among older adults with epilepsy, particularly when integrated with AD biomarkers. Findings support a multimodal framework for assessing AD-related risk in epilepsy and highlight interactive effects of epilepsy chronicity and AD-related processes that can influence cognitive outcomes.

## Introduction

Epilepsy incidence rises in mid- to late-life, contributing to a growing population at increased risk for cognitive impairment^1–3^. Accumulating evidence suggests a bidirectional relationship between epilepsy and Alzheimer’s disease (AD)^4–6^, potentially reflecting overlapping mechanisms of neurodegeneration (e.g., amyloid or tau pathology)^7–11^. Despite intersecting risks across epilepsy, aging, and AD, it remains unclear whether established markers of AD-related neurodegeneration developed in aging cohorts are informative in individuals with epilepsy.

Prior structural neuroimaging studies demonstrate heterogeneous cortical changes in older adults with epilepsy, with evidence of both cortical atrophy and relative cortical preservation. Kaestner et al. (2021) found similar cortical atrophy patterns in medial temporal regions among late-onset temporal lobe epilepsy compared to amnestic mild cognitive impairment (MCI), alongside significant memory and language impairment^12^. Additional studies of late-onset unexplained epilepsy (LOUE), characterized by epilepsy onset in mid- to late-life (e.g., after age 55) without an identifiable structural or other cause, show lower cortical thickness or volumes relative to healthy control groups^13–16^. However, other LOUE studies report relatively preserved gray matter^17^ and hippocampal volumes^17,18^, or potential hypertrophic cortical changes^19^.

Epilepsy can also be associated with widespread gray and white matter atrophy across the lifespan, exceeding typical age-associated changes, suggesting a potentially progressive process that may interact with aging^20^. While many studies on older adults with epilepsy focus on LOUE, individuals with epilepsy onset prior to midlife (early-onset epilepsy [EOE]) may represent a particularly vulnerable subgroup given lifetime seizure burden and other epilepsy-related factors that can contribute to pathological brain and cognitive changes. Given clinical heterogeneity across epilepsy subtypes, it remains difficult to identify subgroups most vulnerable to AD-related neurodegeneration and cognitive decline.

Composite scores of structural MRI metrics in AD-vulnerable brain regions, commonly termed, “AD signatures,” are useful tools in predicting AD-related cognitive decline in aging populations^21–24^. These signatures, typically reflecting cortical thinning or volume loss in temporal regions, were developed and validated across multiple aging cohorts and show links to AD-related biomarkers and risk of progression to MCI or dementia^25–29^. AD signatures are categorized as non-specific markers that become abnormal relatively late on the typical AD continuum, but can be used alongside core measures of amyloid and tau to monitor disease progression and inform prognosis^30^. Application of this validated framework in aging epilepsy populations may allow risk stratification for cognitive decline and inform clinical care in older adults with epilepsy.

In this multicenter cohort, we examined 1) whether older adults with epilepsy mirror AD-related changes, as indexed by a composite AD signature; 2) clinical, cognitive, and biomarker correlates of an AD signature in an epilepsy population; and 3) the utility of an AD signature to predict cognitive impairment among older adults with epilepsy. We leveraged a large, external reference cohort including cognitively unimpaired (CU) older adults, participants with MCI, and those with AD dementia. Notably, our prospectively enrolled sample of older adults with epilepsy allowed for direct examination of differences in AD signatures between those with early and late-onset epilepsy.

## Methods

### Participants

#### Brain Aging and Cognition in Epilepsy (BrACE) cohort

Participants were recruited as part of the BrACE study, a prospective longitudinal multicenter study of cognitive and brain aging in older adults with epilepsy. Inclusion criteria required participants to be ≥ 55 years-old, proficient in English, and diagnosed with focal epilepsy by a board-certified epileptologist according to International League Against Epilepsy criteria^31^. The sample includes both LOUE (onset at or after age 55) and older adults with epilepsy onset earlier than LOUE cohort (referred to here as early-onset epilepsy (EOE). Exclusion criteria included a history of therapeutic epilepsy surgery (e.g., resection or neuromodulation), clinical stroke, space-occupying MRI lesions, large cortical encephalomalacia, or a baseline diagnosis of dementia, primary psychosis, or non-epilepsy-related neurological disease. This study was approved by Institutional Review Boards at the University of California San Diego, Cleveland Clinic, and University of Wisconsin-Madison. Written informed consent was obtained from all participants. A total of 87 BrACE participants (50 with early-onset epilepsy, 37 with late-onset epilepsy) were included in the present analyses.

#### Alzheimer’s Disease Neuroimaging Initiative cohort

Data from the ADNI were used to select comparison samples. The ADNI was launched in 2003 as a public-private partnership, led by Principal Investigator Michael W. Weiner, MD. The primary goal of ADNI has been to test whether biological markers, clinical assessment, and neuropsychological measures can be combined to measure the progression of MCI and early AD. Newly enrolled participants are required to be between the ages of 55-90, and study exclusion criteria included the presence of other neurologic disease (e.g., seizure disorder).

For the current study, participants from ADNI-3 with at least one timepoint with relevant imaging data were selected from three diagnostic groups: cognitively unimpaired (CU), mild cognitive impairment (MCI), and AD dementia. CU participants were defined as retaining a diagnosis of CU at all available timepoints, with no positive amyloid or tau CSF/PET results at any visit (if CSF/PET results were available). Each diagnostic group was separately matched to the full BrACE sample using optimal propensity score matching on age and sex^32^. This yielded matched samples of 128 CU, 128 MCI, and 106 AD participants (Table 1). Data were downloaded from the ADNI database (adni.loni.usc.edu) in January 2026. Informed consent was obtained from all participants, and procedures were approved by the Institutional Review Board of participating institutions.

**Table 1.** BrACE and ADNI sample characteristics.

|  | BrACE<br>Epilepsy<br>*n* = 87 | ADNI<br>CU<br>*n* = 128 | ADNI<br>MCI<br>*n* = 128 | ADNI<br>AD<br>*n* = 106 | Overall group<br>comparison |
| --- | --- | --- | --- | --- | --- |
| Age, mean (SD) | 66.10 (6.86) | 69.19 (4.94) | 69.86 (5.31) | 78.71 (8.05) | $F = 51.49, p < 0.001$ |
| Female sex, *n* (%) | 49 (56.3%) | 77 (60.2%) | 69 (53.9%) | 42 (42.5%) | $\chi^2 = 10.66, p = 0.014$ |
| Education years, mean (SD) | 14.92 (2.53) | 16.56 (2.29) | 15.89 (2.55) | 15.98 (2.52) | $F = 7.84, p < 0.001$ |
| Race, *n* (%) | | | | | $\chi^2 = 10.80, p = 0.767$ |
| American Indian or Alaska Native | 0 (0%) | 1 (<1%) | 0 (0%) | 0 (0%) |  |
| Asian | 1 (1.1%) | 2 (1.6%) | 4 (3.1%) | 1 (<1%) |  |
| Black | 5 (5.7%) | 10 (7.8%) | 8 (6.2%) | 3 (2.8%) |  |
| Multiracial | 3 (3.4%) | 3 (2.3%) | 4 (3.1%) | 2 (1.8%) |  |
| White | 76 (87.4%) | 110 (85.9%) | 111 (86.7%) | 100 (94.3%) |  |
| Unknown | 2 (2.3%) | 2 (1.6%) | 1 (<1%) | 0 (0%) |  |
| Ethnicity, *n* (%) | | | | | $\chi^2 = 10.74, p = 0.097$ |
| Hispanic | 6 (6.9%) | 12 (9.4%) | 4 (3.1%) | 2 (1.9%) |  |
| Non-Hispanic | 81 (93.1%) | 116 (90.6%) | 123 (96.1%) | 104 (98.1%) |  |
| Unknown | 0 (0%) | 0 (0%) | 1 (<1%) | 0 (0%) |  |

ADNI = Alzheimer's Disease Neuroimaging Initiative, BrACE = Brain Aging and Cognition in Epilepsy, CU = cognitively unimpaired, MCI = mild cognitive impairment.

#### Neuropsychological data

BrACE participants completed a comprehensive neuropsychological battery with tests of memory, language, attention/processing speed, and executive function. This cognitive battery was designed to be overlapping with the battery used in ADNI to facilitate comparisons across study cohorts. The International Classification of Cognitive Disorders in Epilepsy (IC-CoDE) was applied to classify cognitive impairment in the BrACE cohort^33,34^. Due to lack of availability of visuospatial tests in the battery, a four-domain classification schema was used (including memory, language, attention/processing speed, and executive function). Participants were classified as cognitively impaired if they demonstrated deficits in one or more cognitive domains as defined by two or more test scores within a domain falling below 1.0 standard deviation based on normative data. The present analyses focused on binary cognitive classification (e.g., impaired vs. unimpaired in one or more domain).

For use in models with continuous predictors, composite variables representing memory, language, attention, and executive functioning were created by converting all normed scores to z-scores and taking the average across tests within each domain (Supplementary Table S1). ADNI participants completed comprehensive neuropsychological testing and were diagnosed with MCI or AD dementia using well-established methods^35^.

#### Plasma biomarkers

In the BrACE cohort, AD-related biomarkers included plasma levels of β-amyloid (Aβ42/40 ratio), phosphorylated tau-217 (pTau217), phosphorylated tau-181 (pTau181), and neurofilament light chain (NfL). These were selected given that they capture amyloid, tau, and neurodegenerative pathology, consistent with current AD frameworks^30,36^. Blood was collected after a minimum of 6 hours of overnight fasting prior to the study appointment. Plasma levels of Aβ40 and Aβ42 were assayed using Neurology 3-Plex A Advantage Kit, and plasma pTau181 was assayed using Advantage V2.1 Assay Kit, both on the Simoa SR-X platform (101995 and 104111, Quanterix; Billerica, MA). NfL was assayed using the R-PLEX Human Neurofilament L Assay from Meso Scale Discovery (K1517XR, MSD; Rockville, MD). pTau217 was assayed using the S-PLEX Human Tau (pT217) Kit from Meso Scale Discovery (K151APFS, MSD; Rockville, MD). All assays were processed according to manufacturers’ instructions. Biomarkers were analyzed as continuous predictors. Due to non-normal distribution of pTau217, pTau181, and NfL levels, each were log transformed prior to use as continuous variables in models. Winsorization at 3 standard deviations above and below the mean was used to avoid the influence of extreme outliers in biomarker data.

### Imaging acquisition and processing

#### BrACE cohort

T1-weighted images were acquired at each BrACE site on a 3T MRI system (UCSD = GE Discovery MR750 MPRAGE, voxel size =1mm isotropic, TR=.006s, TE=2ms, TI=1.06s, flip angle=8°; CC = Siemens Prisma MPRAGE, voxel size = 1mm isotropic, TR=2.3s, TE=2ms, TI=0.9s, flip angle=9°; UWM = GE Signa Premier, SPGR, TR=.007s, TE=2ms, TI=.4s, flip angle = 11°). FreeSurfer v7.3 software was used to obtain cortical thickness estimates and hippocampal volume^37,38^. The Desikan-Killiany atlas was used for regional parcellation^39^. All images underwent visual quality inspection by a board-certified neuroradiologist.

#### ADNI cohort

Processed structural MRI data were downloaded from the ADNI database in January 2026. ADNI-3 data were processed using FreeSurfer v7.2, regional parcellation used the Desikan-Killiany atlas in line with methods used in the BrACE cohort. As detailed in FreeSurfer release notes, recon-all produces the same results as other 7.X releases, facilitating comparison of FreeSurfer data across BrACE and ADNI cohorts.

#### Calculation of AD signature scores

AD signature scores were calculated for BrACE and ADNI participants with available imaging data using established methods^23,27^. The signature comprises a weighted sum of cortical thickness in 7 regions (entorhinal cortex, middle temporal gyrus, banks of the superior temporal sulcus, superior temporal gyrus, isthmus cingulate, lateral orbitofrontal cortex, and medial orbitofrontal cortex) plus hippocampal volume, with separate weights for left and right hemispheres (Figure 1a). Regions and weights were derived from prior work using a data-driven approach to identify atrophy patterns discriminating mild AD from CU individuals in the ADNI-1 cohort^23^. To apply the signature across ADNI-3 and BrACE samples, age and sex effects were regressed from each ROI using linear models fit within the ADNI-3 CU subsample, with estimated total intracranial volume included as an additional covariate for hippocampal ROIs. Coefficients were applied to all participants to generate residuals that reflect deviations from a healthy, age- and sex-adjusted control group. The resulting composite was scaled such that zero reflects the ADNI CU mean, with lower scores indicating lower cortical thickness or hippocampal volume (hereafter referred to as thickness or volume) across AD-sensitive regions relative to the ADNI CU group. In the current analyses, lower scores are interpreted as more AD-like signatures.

**Figure 1.**
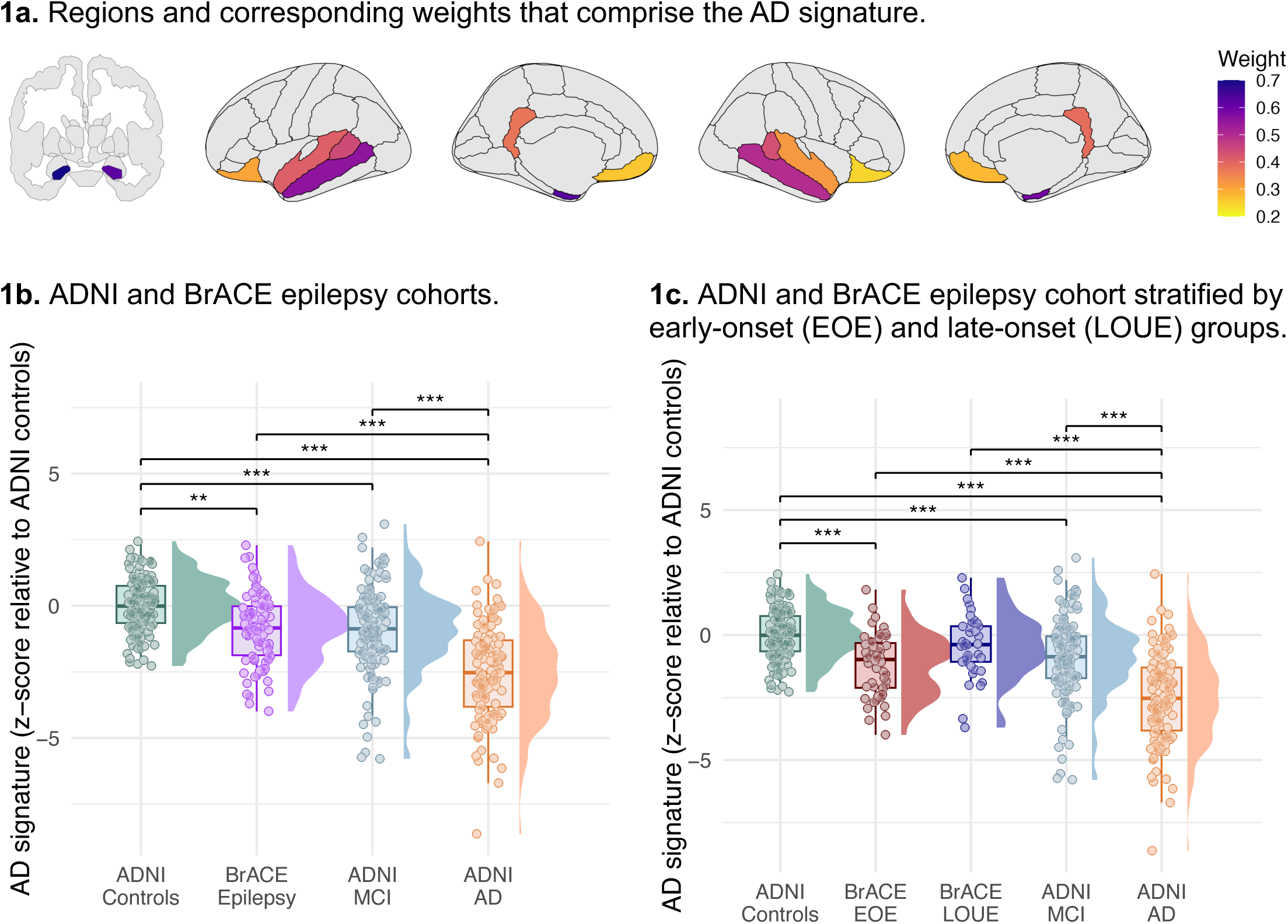
Alzheimer’s disease (AD) signature scores across cohorts. Note: significance levels (prior to adjustment): \**p* < .05, \*\**p* < .01, \*\*\**p* < .001. Partial residuals from models that covaried for age and sex are displayed.

### Statistical analyses

All statistical analyses were conducted in R_v4.4.2_. Linear regression models were fit as described below using the base R *lm()* function. Estimated marginal means and pairwise comparisons were computed using the *emmeans* package, and standardized coefficients were obtained using the *lm.beta* package. Firth penalized likelihood regression, implemented with the *logistf* package, was used for all logistic regression models to reduce small-sample bias and address potential separation issues. To account for multiple comparisons, a false discovery rate (FDR)-corrected threshold of *p* < 0.05 (Benjamini-Hochberg method^40^) was applied separately within each set of analyses: across pairwise group comparisons in aim 1, within each variable domain in aim 2 (demographic/clinical, cognitive, biomarker), and model type in aim 3 (main effects, interaction effects). All models were adjusted for age and sex. Models with cognitive composites or IC-CoDE impairment additionally included years of education as a covariate. Plasma biomarker variables (pTau217, pTau181, Aβ42/40, NfL) were adjusted for sample storage time prior to analysis; history of chronic kidney disease was included as an additional covariate in all biomarker models given its independent influence on plasma biomarker concentrations.

#### 1) AD signatures in BrACE and ADNI cohorts

First, we examined group differences in AD signature scores between the BrACE sample and ADNI comparison groups (ADNI CU, ADNI MCI, ADNI AD). Linear models examined AD signature scores as the outcome with group membership as the primary predictor. Analyses were repeated with BrACE participants stratified by epilepsy onset age (EOE vs. LOUE). To examine whether group differences may be driven by cognitively impaired BrACE participants (*n* = 23), we also repeated analyses excluding this subgroup.

#### 2) Clinical, cognitive, and biomarker correlates with AD signatures in epilepsy

Next, within the BrACE sample, associations between AD signature scores (outcome variable) and a range of clinical, cognitive, and biomarker variables were examined in linear models. Secondary analyses examined potential moderating effects of *APOE*-ε4 status (carriers versus non-carriers) on relationships between clinical, cognitive and biomarker variables with AD signature scores. Due to sparse cell sizes for the variable indicating seizure localization (4 levels), this variable was dichotomized into ‘temporal’ vs. ‘other’ for this set of moderation analyses.

#### 3) Identification of cognitive impairment using AD signatures in epilepsy

Lastly, we tested the utility of the AD signature within the BrACE sample to predict cross-sectional cognitive status (impaired vs. unimpaired) defined by IC-CoDE. To compare against the predictive utility of AD-related plasma biomarkers, logistic regression models were used to test main effects of each plasma biomarker and the AD signature predicting IC-CoDE impairment (coded as 0 for unimpaired and 1 for impaired), with one model specified per predictor of interest. Separate logistic regression models were used to test whether relationships between the AD signature or plasma biomarkers and cognitive impairment differed as a function of epilepsy onset age, with main effects specified under the interaction term (e.g., AD signature*epilepsy onset age; all continuous predictors). Significant interaction effects were further examined by stratifying the sample by early (seizure onset prior to age 55) versus late-onset (seizure onset at or after age 55) epilepsy. Incremental utility of the AD signature was tested by comparing nested logistic regression models (biomarker-only model including pTau217, pTau181, Aβ42/40, NfL; biomarker model plus AD signature) using a penalized likelihood ratio test.

### Sensitivity analyses

Given that the relationships between demographic factors (age, sex) and cortical thickness/volume may differ across ADNI and BrACE samples, we conducted sensitivity analyses with a non-residualized version of the AD signature. This non-residualized signature reflects the same regions and weights described above, with estimated intracranial volume regressed out of hippocampal volume ROIs, but without regressing out age and sex effects based on the ADNI CU sample, which may not generalize to an epilepsy sample. Associations between this non-residualized signature and clinical, cognitive, and biomarker variables within the BrACE sample were examined and are reported in Supplementary Materials.

A second set of sensitivity analyses examined whether broader summary brain measures show similar associations with variables of interest compared to the AD signature. Within an epilepsy sample, an AD signature may simply reflect widespread cortical thinning or hippocampal atrophy that is not tied to other AD-related regions. To test whether global summary measures, such as total cortical thickness or hippocampal volume alone, showed similar effects compared to the AD signature, we repeated all analyses using: 1) average bilateral cortical thickness (across all ROIs, including those comprising the AD signature), and 2) average bilateral hippocampal volume. These models used the same covariates as the main analyses, with estimated total intracranial volume added as an additional covariate for models with hippocampal volume. Effect sizes from these separate models were then compared against those from the main AD signature models.

## Results

### Participant demographics

Demographic information is displayed in Table 1. The ADNI sample was selected to match the BrACE cohort as closely as possible in age and sex; however, the ADNI CU, MCI, and AD groups were older than the BrACE cohort (all pairwise *p*s_adj_ < 0.001). This difference reflects limited overlap in age distributions across the BrACE and ADNI cohorts. Sex was balanced across ADNI CU, MCI, and BrACE groups, though the ADNI AD group showed a lower proportion of female participants relative to the ADNI CU group (pairwise *p*_adj_ = 0.015). The ADNI sample also showed greater years of education relative to the BrACE cohort. Table 2 displays sample characteristics in the BrACE cohort stratified by EOE and LOUE groups.

**Table 2.**
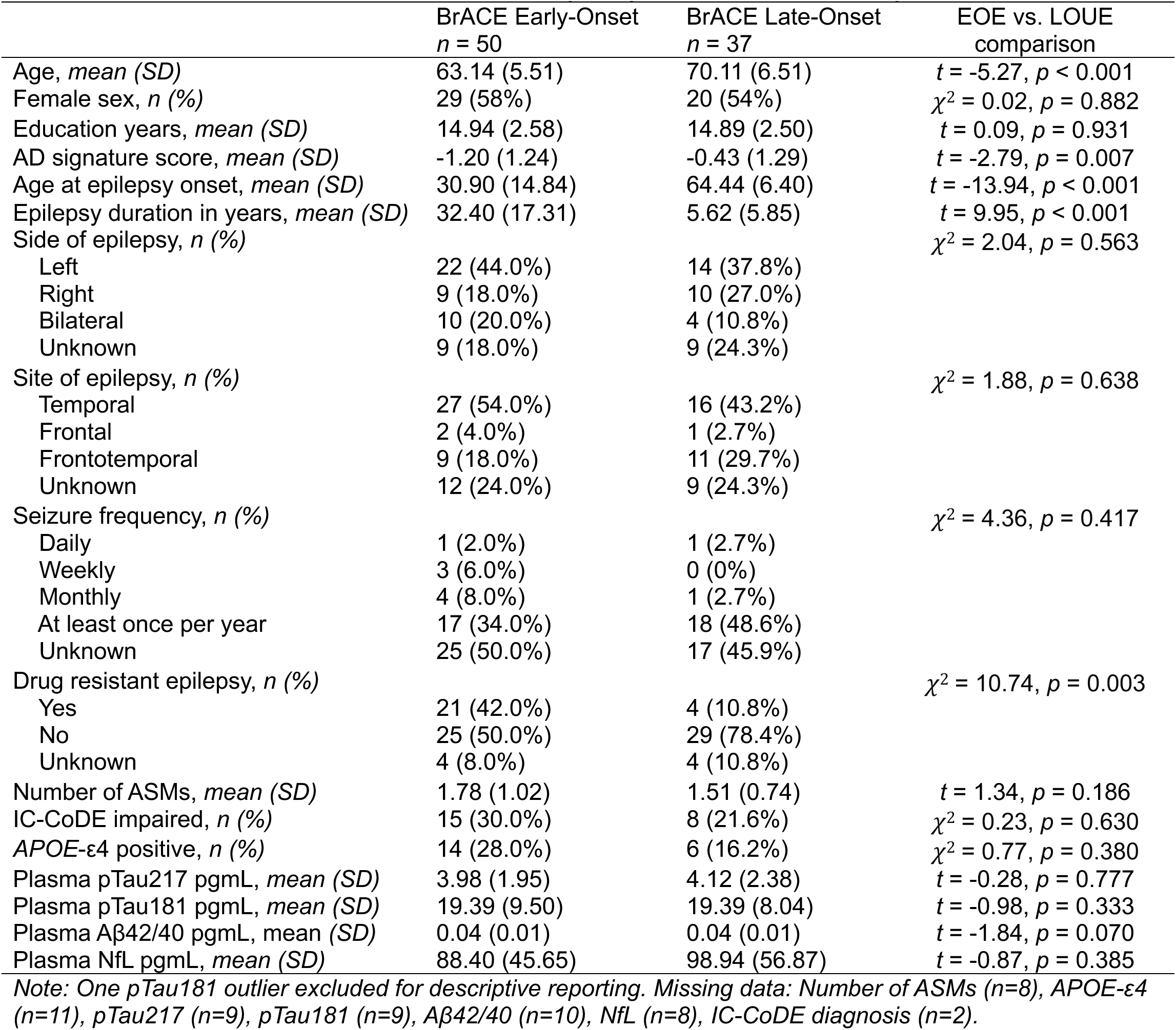
BrACE sample characteristics stratified by early-vs. late-onset epilepsy.

#### 1) AD signatures in BrACE and ADNI cohorts

AD signature scores differed significantly by group (F = 56.6, *p* < 0.001). The BrACE cohort showed more AD-like signature scores compared to the ADNI CU group (β= −0.43, pairwise *p*_adj_ < 0.001), reflecting lower cortical thickness or volume across AD-related regions (Figure 1b). The BrACE cohort showed similar AD signature scores compared to the ADNI MCI group (β = 0.10, pairwise *p*_adj_ = 0.384) and less AD-like signatures compared to the ADNI AD group (β = 1.22, pairwise *p*_adj_ < 0.001). As expected, given the design of the signature, the ADNI MCI and AD groups showed more AD-like signature scores relative to the ADNI CU group (βs= −0.53 to −1.64, pairwise *p*s_adj_ < 0.001).

Stratifying the BrACE sample by EOE and LOUE revealed that those with EOE (β= −0.57, pairwise *p*_adj_ < 0.001), but not LOUE (β= −0.26, pairwise *p*_adj_ = 0.102), showed more AD-like signature scores relative to the ADNI CU group (Figure 1c). Results were similar when restricting the BrACE sample to only CU participants (n = 64), such that the BrACE CU sample still showed more AD-like signature scores relative to the ADNI CU sample (β= −0.39, pairwise *p*_adj_ = 0.003). This pattern similarly was stronger for the EOE (β= −0.46, pairwise *p*_adj_ = 0.006) compared to the LOUE (β= −0.32, pairwise *p*_adj_ = 0.062) group. Average cortical thickness in AD signature regions across EOE and LOUE groups is displayed in Supplementary Figure S1.

#### 2) Clinical, cognitive, and biomarker correlates with AD signatures in epilepsy

In models examining main effects, worse memory performance (β = 0.30, *p*_adj_ = 0.006) and higher levels of NfL (β = −0.30, *p*_adj_ = 0.050) were associated with more AD-like signatures, Figure 2a). There were no significant main effects of epilepsy-related clinical variables, and there were no significant correlations between the AD signature and amyloid or tau biomarkers in the BrACE cohort. Table 3 displays full results from these models.

**Figure 2.**
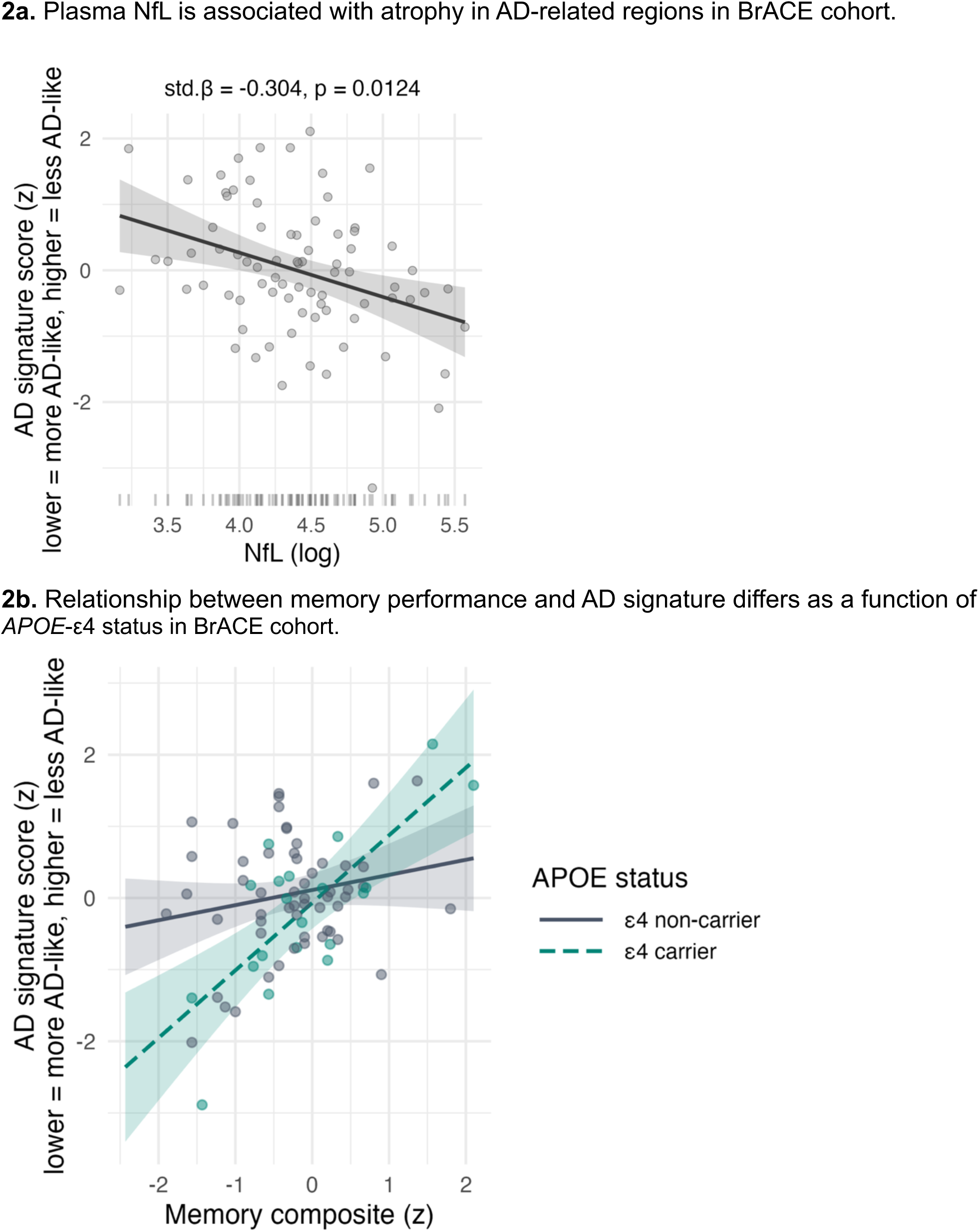
Biomarker and cognitive correlates with AD signature in older adults with epilepsy. **2a.** Plasma NfL is associated with atrophy in AD-related regions in BrACE cohort. **2b.** Relationship between memory performance and AD signature differs as a function of *APOE*-ε4 status in BrACE cohort.

**Table 3.** Clinical, cognitive, and biomarker associations with AD signature scores in full BrACE sample.

| | $\beta$ | Partial $R^2$ | $p_{adj}$ |
| --- | --- | --- | --- |
| **Clinical characteristics** |  |  |  |
| Seizure localization | -- | 0.119 | 0.161 |
| Days since last seizure | -0.193 | 0.046 | 0.308 |
| Years of education | 0.109 | 0.016 | 0.503 |
| Duration of epilepsy | -0.107 | 0.015 | 0.503 |
| Side of seizure focus | -- | 0.045 | 0.503 |
| APOE-$\epsilon 4$ status | -0.088 | 0.010 | 0.503 |
| Drug resistance status | -- | 0.024 | 0.503 |
| Early vs. late onset | 0.074 | 0.005 | 0.566 |
| Number of ASMs | 0.045 | 0.002 | 0.666 |
| **Cognitive performances** |  |  |  |
| Memory composite | 0.304 | 0.117 | 0.006* |
| Language composite | 0.180 | 0.042 | 0.122 |
| Attention composite | -0.071 | 0.007 | 0.615 |
| Executive function composite | 0.002 | < 0.001 | 0.987 |
| **Plasma AD biomarker levels** |  |  |  |
| NfL (log) | -0.304 | 0.083 | 0.050* |
| pTau181 (log) | -0.151 | 0.025 | 0.364 |
| Aβ42/40 ratio | 0.083 | 0.010 | 0.549 |
| pTau217 (log) | 0.034 | 0.001 | 0.777 |

*Effect sizes are reported as standardized regression coefficients ( $\beta$ ) for continuous and binary predictors, and partial $R^2$ for all predictors including categorical variables with >2 levels.*

In secondary interaction models, the relationship between memory performance and AD signature scores differed as a function of *APOE*-ε4 status (interaction *p*_adj_ = 0.023; Figure 2b). Individuals with an *APOE*-ε4 allele showed a stronger relationship between memory performance and AD signature score (β = 0.72) compared to those without an *APOE*-ε4 allele (β = 0.16). There were no other significant moderating effects of *APOE*-ε4 status after correction for multiple comparisons.

#### 3) Identification of cognitive impairment using AD signatures in epilepsy

In the full sample of BrACE participants, main effects for each plasma biomarker and the AD signature predicting cross-sectional cognitive impairment were nonsignificant (*p*s_adj_ > 0.05). However, interaction models revealed that more AD-like signatures were associated with cognitive impairment specifically among those with earlier epilepsy onset (OR = 0.53, *p* = 0.047) compared to later epilepsy onset (OR = 1.10, *p* = 0.852; overall interaction *p*_adj_ = 0.043, Figure 3). The relationship between pTau217 and cognitive impairment showed a similar pattern, such that higher levels of pTau217 were associated with cognitive impairment specifically among those with earlier epilepsy onset (OR = 3.24, *p* = 0.003) compared to those with later epilepsy onset (OR = 0.50, *p* = 0.263; overall interaction *p*_adj_ = 0.043).

**Figure 3.**
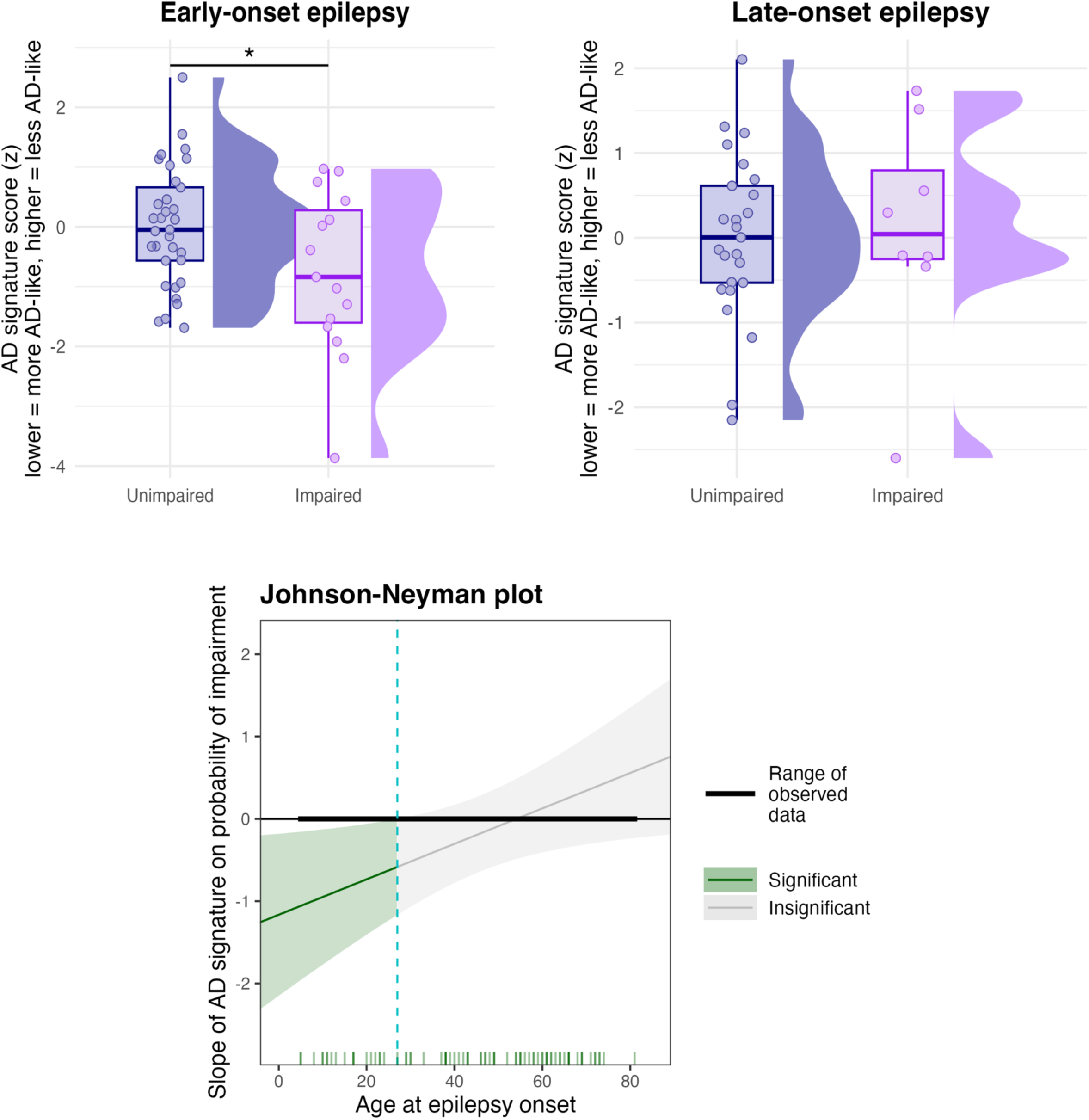
AD signature is associated with cognitive impairment among older adults with early-onset epilepsy, but not among those with late-onset epilepsy.

Among those with EOE, adding the AD signature to a model with plasma biomarkers significantly improved prediction of cognitive impairment (χ^2^ = 4.18, *p* = 0.041). Adding the AD signature to a model with plasma biomarkers among those with LOUE did not improve predictions. Relationships between other plasma biomarkers (pTau181, Aβ42/40, NfL) and cognitive impairment did not differ by epilepsy onset age. Analyses were repeated using years of epilepsy duration as the moderator variable, rather than age of epilepsy onset, and results remained similar (Supplementary Figure S2).

### Sensitivity analyses

We conducted sensitivity analyses with a non-residualized version of the AD signature, as age and sex effects based on the ADNI CU sample may not generalize to an epilepsy sample. Overall, the pattern of results remained similar (Supplementary Table S2), suggesting that findings described above were not otherwise explained by potential statistical artifacts that can arise when applying age- or sex-derived slopes from a healthy control sample to a different patient population.

In a second set of sensitivity analyses, we examined whether global summary measures, such as total cortical thickness or hippocampal volume alone, showed similar effect sizes compared to the AD signature. Within the BrACE cohort, average global cortical thickness was positively associated with memory performance (β = 0.31, *p*_adj_ = 0.020), showing a similar effect size compared to the AD signature (β = 0.30). However, within BrACE there were no significant relationships between average cortical thickness and biomarker levels. Average cortical thickness was not significantly associated with IC-CoDE impairment across any level of epilepsy onset age or duration (all interaction *p*s > 0.05). Further, adding average cortical thickness to a model with plasma biomarkers did not improve prediction of IC-CoDE impairment in EOE or LOUE. Similarly, there were no significant relationships between average hippocampal volume and clinical, cognitive, or biomarker levels after adjustment for multiple comparisons. Average hippocampal volume did not predict IC-CoDE impairment across any level of epilepsy onset age or duration, and did not improve prediction of IC-CoDE impairment in EOE or LOUE beyond plasma biomarker levels. Full results from these sensitivity analyses are reported in Supplementary Tables S3-S4.

## Discussion

This multicenter study demonstrates that an AD neuroimaging signature could serve as a useful tool for identifying clinically relevant subgroups among older adults with epilepsy, particularly when used alongside other AD biomarkers. Older adults with epilepsy showed more AD-like signatures relative to a healthy control sample, reflecting lower cortical thickness or volume across a set of AD-related brain regions. Within the epilepsy cohort, this AD signature was associated with plasma NfL, memory performance, and predicted greater odds of cognitive impairment specifically among those with EOE but not LOUE. Among those with EOE, the AD signature significantly improved identification of cognitive impairment over and beyond the effects of plasma biomarkers, underscoring the importance of multimodal approaches to understanding AD-related risk within this population.

The BrACE sample showed more AD-like signatures compared to a healthy control sample, despite being younger. Interestingly, this pattern was stronger among those with EOE (β= −0.57) compared to those with LOUE (β= −0.26). This is in line with recent work by our group in the same BrACE sample that demonstrated those with EOE showed significantly higher odds of AD biomarker abnormality compared to those with LOUE, despite the EOE group being an average of 6.5 years younger^41^. These findings may suggest the role of cumulative seizure burden, epilepsy-related factors, and vascular risk factors across the lifespan that can interact with AD-related processes to influence neurodegenerative outcomes. Prior work has shown that EOE can be associated with increased brain amyloid levels in late middle age, particularly among *APOE*-ε4 carriers^42^, and greater progressive amyloid accumulation relative to healthy control samples^43^. *APOE*-ε4 status has additionally been shown to interact with epilepsy duration to influence cognitive outcomes, such that *APOE*-ε4 carriers with EOE or longstanding seizure history demonstrate worse cognitive performance or more rapid cognitive decline over time^44,45^. Secondary analyses in the present study revealed that individuals with an *APOE*-ε4 allele showed a stronger relationship between memory performance and AD signature scores compared to those without an *APOE*-ε4 allele. Collectively, these findings highlight the importance of considering epilepsy chronicity and genetic vulnerability when examining brain structure-cognition relationships in older adults with epilepsy and suggest that certain subgroups may be particularly susceptible to overlapping or synergistic pathological processes.

Existing structural MRI studies in older adults with epilepsy have yielded mixed findings, likely reflecting clinical heterogeneity in epilepsy populations. Studies focused on LOUE demonstrate both cortical thinning in AD-related regions relative to healthy controls^12–16^ as well as relative cortical preservation or potential hypertrophic changes compared to control groups^17–19^. Few studies have directly compared cortical atrophy patterns between EOE and LOUE groups. In a separate cohort of older adults with drug-resistant epilepsy (DRE), LOUE was associated with more prominent cortical thinning in AD-related regions compared to EOE^12^, whereas our current results suggest that those with EOE show more robust atrophy patterns as indexed by an AD signature compared to LOUE. These discrepant findings may partly reflect differences in clinical characteristics, particularly DRE prevalence. In the present study, most participants had relatively well controlled epilepsy, though DRE rates were higher in EOE than LOUE (42% vs. 11%) consistent with evidence that DRE negatively impacts brain aging^46^ and can be associated with AD neuropathologic change^9^.

The 2024 Alzheimer’s Association research criteria for the diagnosis and staging of AD^30^ suggests that structural MRI measures, such as AD signatures, are non-specific markers of AD pathology that become abnormal relatively late on the AD continuum and are closely tied to the onset of cognitive impairment. The 2024 International Working Group Recommendations^36^ importantly detail that AD biomarkers on their own are not sufficient indicators of whether an individual will develop cognitive impairment. Evidence from aging cohorts shows that the risk of developing MCI or dementia is increased among individuals with abnormal biomarker profiles, though there is considerable variability in clinical progression rates^36,47^. Our current results revealed that the AD signature and plasma pTau217 were associated with cognitive impairment specifically among those with EOE, suggesting cumulative effects of disease duration that may contribute to cognitive vulnerability in the presence of AD-related risk. Further, combining plasma AD biomarkers and an AD signature better identified a subgroup with greater odds of cognitive impairment than using any measure on its own, highlighting the importance of contextualizing AD-related risk with multiple indicators in populations with epilepsy to more accurately identify individuals acutely at risk for cognitive decline.

Whether the overall pattern of cortical atrophy captured by the AD signature reflects AD-related, epilepsy-related, or a combination of these processes in older adults with epilepsy remains difficult to ascertain. Interestingly, the AD signature was associated with plasma levels of NfL, but not with plasma markers of amyloid or tau in the BrACE sample. This may suggest that atrophy patterns resembling AD in older adults with epilepsy may be more closely tied to nonspecific neurodegeneration rather than amyloid or tau-driven AD-specific processes. Sensitivity analyses demonstrated that the AD signature showed a stronger relationship with plasma NfL and better identified subgroups with cognitive impairment among those with EOE, compared to average cortical thickness or hippocampal volume alone, suggesting that atrophy patterns reflected in the AD signature contribute meaningful information even when not directly associated with amyloid or tau biomarkers. These results support the utility of an AD signature as a complementary marker in older adults with epilepsy, particularly those with earlier disease onset and longer disease duration.

It is important to note that processes underlying cognitive impairment in epilepsy are heterogeneous and often include cerebrovascular disease^3,5,48–50^, which is not overtly reflected in the cortical AD signature. Thus, our results also highlight the limitations of applying AD-derived frameworks directly to epilepsy populations. Structural signatures captured meaningful variation beyond global cortical thinning or hippocampal volume alone, but they do not fully account for other contributors to cognitive impairment in epilepsy, including vascular pathology and network-level dysfunction. Incorporating white matter microstructure, functional imaging, and cerebrovascular integrity measures could therefore strengthen this approach by capturing a broader range of relevant pathology in epilepsy populations.

These findings should be interpreted in light of several limitations. First, the AD signature that we used was developed in the ADNI-1 sample in prior work based on its utility to distinguish mild AD from healthy controls. It is possible that an AD signature designed specifically for epilepsy populations with metrics beyond cortical thickness or volume may yield optimal results for distinguishing risk for AD-related cognitive decline in this cohort, and should be explored in future work. Some of the same ADNI-1 participants that were used in the development of the signature may have contributed data at subsequent ADNI visits in the present analyses if they met our inclusion criteria, though only data from ADNI-3 were used in the current study. Regarding generalizability of results, less than one-third of the BrACE sample showed cognitive impairment defined by IC-CoDE, which may restrict generalizability to epilepsy populations showing greater levels of cognitive impairment. Further, the BrACE cohort represented a sample of patients with relatively well controlled epilepsy, and as such, these findings may not generalize to patients with more severe forms of epilepsy who may be at a greater risk of brain and cognitive deterioration. Finally, current results are based on cross-sectional data; the utility of an AD signature to predict longitudinal cognitive outcomes in older adults with epilepsy should be explored in future work.

In summary, our findings demonstrate the utility of composite MRI measures of AD-related neurodegeneration as part of a multimodal framework alongside plasma AD biomarkers to identify clinically relevant subgroups in epilepsy populations. Relationships across memory performance and levels of plasma NfL underscore the relevance of an AD signature among older adults with epilepsy, and findings among those with EOE highlight interactive effects of epilepsy chronicity and AD-related processes that can influence cognitive outcomes. Adaptation of existing neurodegenerative models to epilepsy populations may improve early identification and guide targeted monitoring and intervention strategies in older adults with epilepsy at elevated risk for cognitive decline.

## Data Availability

The ADNI data analyzed in the current study are available in the ADNI database (adni.loni.usc.edu). The BrACE data supporting the findings of this study are available on request from the senior author (C.R.M.).

## Acknowledgement

This research was supported by the National Institutes of Health R01NS120976. A.R. is supported by NINDS 1K23NS138682-01A1. R.B. also receives support from NINDS R01NS135080, U54NS092090, and R33AG039729 and the Cleveland Clinic Epilepsy Center. AS is supported by NINDS K23 and the Alzheimer’s Association (NIAP25-1438957). IZ is supported by the American Epilepsy Society (Award ID: 1067206), Alzheimer’s Association (Grant: AACSFD-22-974008), and NIH/NIA (1K23AG084893). IW receives support from NINDS 2R01 NS109439.

Data used in preparation of this article were obtained from the ADNI database (adni.loni.usc.edu). As such, the investigators within the ADNI contributed to the design and implementation of ADNI and/or provided data but did not participate in analysis or writing of this report. A complete listing of ADNI investigators can be found at: http://adni.loni.usc.edu/wp-content/uploads/how_to_apply/ADNI_Acknowledgement_List.pdf.

Data collection and sharing for the Alzheimer’s Disease Neuroimaging Initiative (ADNI) is funded by the National Institute on Aging (National Institutes of Health Grant U19 AG024904). The grantee organization is the Northern California Institute for Research and Education.

In the past, ADNI has also received funding from the National Institute of Biomedical Imaging and Bioengineering, the Canadian Institutes of Health Research, and private sector contributions through the Foundation for the National Institutes of Health (FNIH) including generous contributions from the following: AbbVie, Alzheimer’s Association; Alzheimer’s Drug Discovery Foundation; Araclon Biotech; BioClinica, Inc.; Biogen; Bristol-Myers Squibb Company; CereSpir, Inc.; Cogstate; Eisai Inc.; Elan Pharmaceuticals, Inc.; Eli Lilly and Company; EuroImmun; F. Hoffmann-La Roche Ltd and its affiliated company Genentech, Inc.; Fujirebio; GE Healthcare; IXICO Ltd.; Janssen Alzheimer Immunotherapy Research & Development, LLC.; Johnson & Johnson Pharmaceutical Research & Development LLC.; Lumosity; Lundbeck; Merck & Co., Inc.; Meso Scale Diagnostics, LLC.; NeuroRx Research; Neurotrack Technologies; Novartis Pharmaceuticals Corporation; Pfizer Inc.; Piramal Imaging; Servier; Takeda Pharmaceutical Company; and Transition Therapeutics.

