## Supplementary materials for "Alzheimer’s disease neuroimaging signature aids identification of cognitive impairment in older adults with early-onset epilepsy"

**Table S1.** Neuropsychological tests and normative data used for IC-CoDE diagnoses.

| Test | IC-CoDE domain | Normative data |
|---|---|---|
| Rey Auditory Verbal Learning Test (RAVLT) Delayed Recall | Memory | Mayo Older Americans Normative Study[1] |
| Wechsler Memory Scale 4th Edition (WMS-IV) Logical Memory (LM) Story B Delayed Recall | Memory | Mayo Older Americans Normative Study[1] |
| WMS-IV Visual Reproduction (VR) Delayed Recall | Memory | WMS-IV Manual[2] |
| Multilingual Naming Test (MINT) | Language | Uniform Data Set (UDS)[3] |
| Auditory Naming Test (ANT) Total Score | Language | Hamberger et al. (2022)[4] |
| Semantic Fluency | Language | Expanded Halstead-Reitan Battery[5] |
| Trail-Making Test Condition A (TMT-A) | Processing Speed/ Attention | Expanded Halstead-Reitan Battery[5] |
| Alzheimer's Disease Assessment Scale–Cognitive Subscale (ADAS-Cog) Number Cancellation Subtest | Processing Speed/ Attention | ADNI Normal Control Sample[6] |
| Letter Fluency | Executive Function | Expanded Halstead-Reitan Battery[5] |
| Trail-Making Test Condition B (TMT-B) | Executive Function | Expanded Halstead-Reitan Battery[5] |

[1] Ivnik RJ, Malec JF, Smith GE, Tangalos EG, Petersen RC, Kokmen E, Kurland LT. Mayo's older Americans normative studies: updated AVLT norms for ages 56 to 97. The Clinical Neuropsychologist. 1992;6(S1):83-104.
[2] Wechsler D. WMS-IV: Wechsler memory scale: PsychCorp; 2009.
[3] Weintraub S, Besser L, Dodge HH, Teylan M, Ferris S, Goldstein FC, et al. Version 3 of the Alzheimer Disease Centers' neuropsychological test battery in the Uniform Data Set (UDS). Alzheimer disease and associated disorders. 2018;32(1):10.
[4] Hamberger MJ, Heydari N, Caccappolo E, Seidel WT. Naming in older adults: Complementary auditory and visual assessment. Journal of the International Neuropsychological Society. 2022;28(6):574-87.
[5] Heaton R, Miller SW, Taylor MJ, Grant-Isibor I. Revised comprehensive norms for an expanded Halstead-Reitan Battery: Demographically adjusted neuropsychological norms for African American and Caucasian adults. 2004.
[6] Cancellation raw scores were converted into z-scores based on data from a sample of 370 cognitively normal older adults from the Alzheimer's Disease Neuroimaging Initiative (ADNI) database (adni.loni.usc.edu).

**Table S2. Clinical, cognitive, and biomarker associations with non-residualized AD signature scores in full BrACE sample.**

| | **β** | **Partial $R^2$** | **$p_{adj}$** |
|---|---|---|---|
| **Clinical characteristics** | | | |
| Epilepsy localization | -- | 0.124 | 0.131 |
| Days since last seizure | -0.215 | 0.046 | 0.307 |
| Years of education | 0.123 | 0.015 | 0.492 |
| Duration of epilepsy | -0.126 | 0.016 | 0.492 |
| Side of epilepsy focus | -- | 0.047 | 0.492 |
| *APOE*-ε4 status | -0.104 | 0.011 | 0.499 |
| Drug resistance status | -- | 0.024 | 0.499 |
| Early vs. late onset | 0.088 | 0.006 | 0.549 |
| Number of ASMs | 0.056 | 0.003 | 0.633 |
| **Cognitive performances** | | | |
| Memory composite | 0.347 | 0.116 | 0.006* |
| Language composite | 0.208 | 0.043 | 0.117 |
| Attention composite | -0.081 | 0.006 | 0.623 |
| Executive function composite | 0.004 | < 0.001 | 0.968 |
| **Plasma AD biomarker levels** | | | |
| NfL (log) | -0.347 | 0.081 | 0.052 |
| pTau181 (log) | -0.170 | 0.024 | 0.380 |
| Aβ42/40 ratio | 0.096 | 0.009 | 0.554 |
| pTau217 (log) | 0.040 | 0.001 | 0.769 |

*Effect sizes are reported as standardized regression coefficients (β) for continuous and binary predictors, and partial $R^2$ for all predictors including categorical variables with >2 levels.*

**Table S3. Clinical, cognitive, and biomarker associations with average bilateral cortical thickness in full BrACE sample.**

| | **β** | **Partial $R^2$** | $\boldsymbol{p}_{adj}$ |
|---|---|---|---|
| **Clinical characteristics** | | | |
| Epilepsy localization | -- | 0.126 | 0.117 |
| Days since last seizure | -0.139 | 0.019 | 0.410 |
| Years of education | 0.113 | 0.013 | 0.410 |
| Duration of epilepsy | -0.139 | 0.019 | 0.410 |
| Side of epilepsy focus | -- | 0.067 | 0.410 |
| *APOE*-ε4 status | -0.115 | 0.013 | 0.410 |
| Drug resistance status | -- | 0.026 | 0.411 |
| Early vs. late onset | 0.130 | 0.012 | 0.410 |
| Number of ASMs | 0.033 | 0.001 | 0.776 |
| **Cognitive performances** | | | |
| Memory composite | 0.310 | 0.092 | 0.020* |
| Language composite | 0.135 | 0.018 | 0.449 |
| Attention composite | -0.104 | 0.011 | 0.467 |
| Executive function composite | 0.014 | < 0.001 | 0.901 |
| **Plasma AD biomarker levels** | | | |
| NfL (log) | -0.238 | 0.039 | 0.361 |
| pTau181 (log) | -0.026 | 0.001 | 0.844 |
| Aβ42/40 ratio | 0.026 | 0.001 | 0.844 |
| pTau217 | 0.094 | 0.006 | 0.844 |

*Effect sizes are reported as standardized regression coefficients (β) for continuous and binary predictors, and partial $R^2$ for all predictors including categorical variables with >2 levels.*

**Table S4. Clinical, cognitive, and biomarker associations with average bilateral hippocampal volume in full BrACE sample.**

| | **β** | **Partial $R^2$** | $p_{adj}$ |
|---|---|---|---|
| **Clinical characteristics** | | | |
| Epilepsy localization | -- | 0.008 | 0.940 |
| Days since last seizure | -0.036 | 0.002 | 0.940 |
| Years of education | 0.083 | 0.010 | 0.940 |
| Duration of epilepsy | -0.067 | 0.006 | 0.940 |
| Side of epilepsy focus | -- | 0.005 | 0.940 |
| *APOE*-ε4 status | -0.109 | 0.019 | 0.940 |
| Drug resistance status | -- | 0.007 | 0.940 |
| Early vs. late onset | 0.016 | 0.001 | 0.940 |
| Number of ASMs | -0.078 | 0.009 | 0.940 |
| **Cognitive performances** | | | |
| Memory composite | 0.085 | 0.011 | 0.710 |
| Language composite | 0.171 | 0.043 | 0.240 |
| Attention composite | < 0.001 | < 0.001 | 0.998 |
| Executive function composite | 0.003 | < 0.001 | 0.998 |
| **Plasma AD biomarker levels** | | | |
| NfL (log) | -0.221 | 0.054 | 0.187 |
| pTau181 (log) | -0.127 | 0.022 | 0.283 |
| Aβ42/40 ratio | 0.124 | 0.024 | 0.283 |
| pTau217 (log) | -0.026 | 0.001 | 0.806 |

*Effect sizes are reported as standardized regression coefficients (β) for continuous and binary predictors, and partial $R^2$ for all predictors including categorical variables with >2 levels.*

**Figure S1. Cortical thickness across AD signature regions in BrACE EOE and LOUE.** Z-scores represent age- and sex-adjusted cortical thickness values relative to the ADNI CU group.

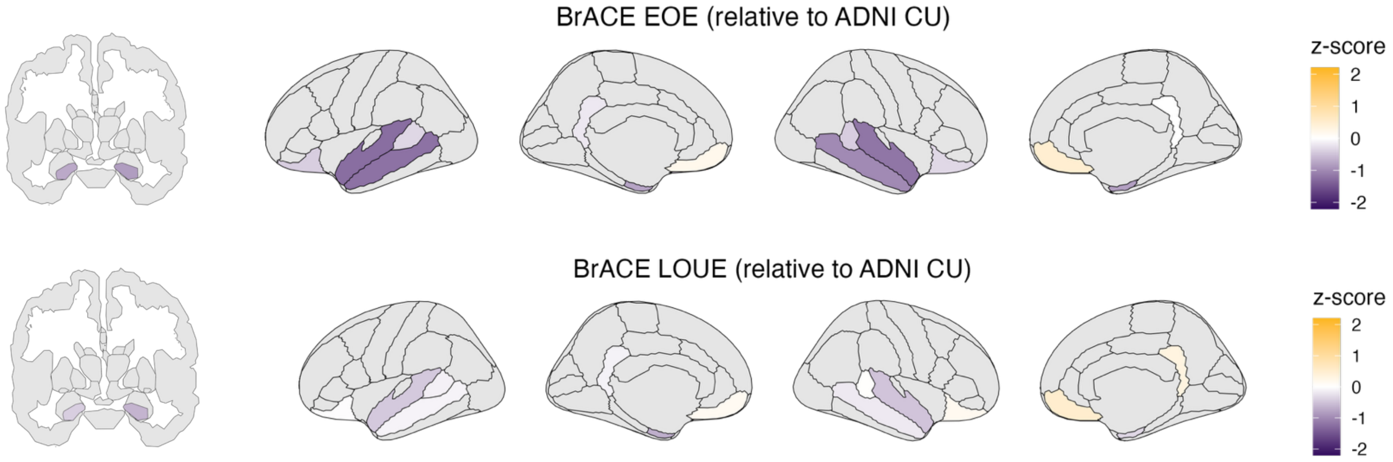

**Identification of cognitive impairment using AD signatures in epilepsy: Stratification by epilepsy duration rather than epilepsy onset age**

Analyses in section 3 of the main text were repeated using epilepsy duration rather than epilepsy onset age. In line with results reported in the main text, interaction models revealed that more AD-like signatures were associated with cognitive impairment specifically among those with longer epilepsy duration compared to shorter duration (overall interaction $p_{adj}$ = 0.027; Figure S2). The relationship between pTau217 and cognitive impairment showed a similar pattern, such that higher levels of pTau217 were associated with cognitive impairment specifically among those with longer compared to shorter epilepsy duration, though this interaction effect did not survive correction for multiple comparisons (overall interaction $p_{adj}$ = 0.068).

Among those with longer epilepsy duration (defined using median split at 20 years of epilepsy duration), adding the AD signature to a model with plasma biomarkers significantly improved prediction of cognitive impairment ($\chi^2$ = 4.73, $p$ = 0.029). Adding the AD signature to a model with plasma biomarkers among those with shorter epilepsy duration did not improve predictions. Relationships between other plasma biomarkers (pTau181, Aβ42/40, NfL) and cognitive impairment did not differ by epilepsy duration.

**Figure S2.** Johnson-Neyman plot depicting significant moderating effect of epilepsy duration on the relationship between AD signature and cognitive impairment.

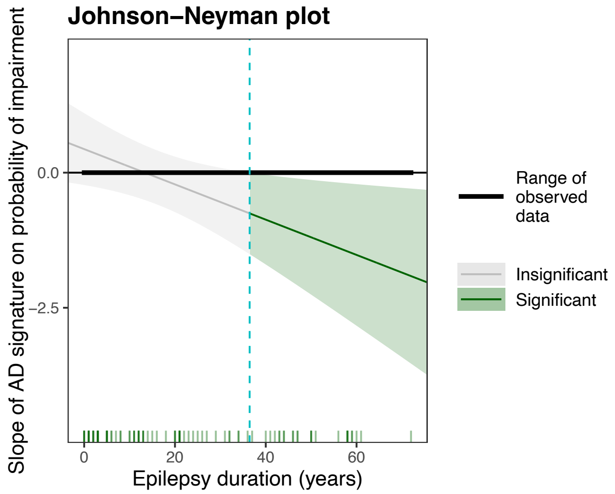
